# Effectiveness of quarantine and testing to prevent COVID-19 transmission from arriving travelers

**DOI:** 10.1101/2020.11.02.20224568

**Authors:** W. Alton Russell, David L. Buckeridge

**Affiliations:** MGH Institute for Technology Assessment, Harvard Medical School, Boston, MS, USA; School of Population and Global Health, McGill University, Montreal, QC, Canada

## Abstract

**Objective:** To assess the efficacy of policies designed to reduce the risk of international travelers importing SARS-CoV-2 into a country.

**Method:** We developed a simulation model and compared mandatory quarantine, testing, and combined quarantine and testing. We assessed the sensitivity of policy effectiveness to the timing of testing, compliance with quarantine and isolation, and other factors.

**Results:** In the base scenario, a 2-day quarantine reduced more risk than testing alone. The effectiveness of a 5-day quarantine requirement with perfect compliance was similar to a 14-day quarantine with moderate compliance. Testing 72h before arrival reduced less than 10% of in-country transmission risk across all scenarios. The addition of testing to quarantine added value for shorter quarantine lengths, when testing compliance was enforced, and when testing was performed near the end of quarantine.

**Conclusions:** Quarantine is more effective at preventing SARS-CoV-2 transmission from arriving travelers than testing alone, but testing combined with quarantine can add value if longer quarantine requirements are infeasible. Enforcing compliance with quarantine and isolation is critical. Requiring a negative test up to 72h before arrival may have limited effectiveness.

## INTRODUCTION

Many countries have implemented policies to reduce the risk of arriving travelers introducing new SARS-CoV-2 transmission chains, particularly for new genetic variants of the virus. Many factors that may influence the relative effectiveness of these policies, including quarantine compliance and the timing of infection relative to arrival, have not been assessed. Additionally, the effectiveness of requiring a negative SARS-CoV-2 test up to 72-hours before arrival, recently implemented for international travelers arriving to the United States, is unknown. We developed a simulation model to estimate the effectiveness of mandatory quarantine, SARS-CoV-2 testing, and combined quarantine and testing for arriving travelers and analyzed the influence of many factors. We also developed a flexible webtool for evaluating policies in context-specific scenarios.

## METHODS

We simulated the infection phases for travelers with asymptomatic infections (‘pre-infectious’ and ‘infectious’) and with symptomatic infections (‘pre-infectious,’ ‘pre-symptomatic-infectious,’ and ‘symptomatic-infectious’). For each traveler, we calculated the number of days at risk of community transmission (i.e., infectious and not in quarantine or isolation) under each policy. We calculated the ‘adjusted’ days at-risk of community transmission by applying a weight to reflect a lower transmission risk for travelers with asymptomatic infections [1]. We calculated the percent risk reduced by each policy as compared to a ‘no intervention’ scenario. These metrics were chosen because they are independent of the prevalence of SARS-CoV-2 infection. We also developed a webtool that allows users to calculate context-specific outcomes based on local data or assumptions (https://shiny.mchi.mcgill.ca/arussel/quarantineTesting/). For all metrics, we reported the median and a 98% credible interval (CrI) that reflects uncertainty in the distribution of infection phase durations.

We simulated mandatory quarantine policies of 0 – 14 days and four testing policies: testing 72h before arrival, testing on arrival (with and without enforced isolation following a positive test), and testing 24h before quarantine end. In the ‘24h before quarantine end’ scenario, travelers who did not comply with quarantine were not tested. Travelers who tested positive in the ‘72h before arrival’ and ‘on arrival with enforcement’ scenarios were assumed to never enter the community. In the base scenario, we assumed 40% of infections were asymptomatic, 80% of travelers would comply with quarantine, 80% would isolate when symptomatic, 90% would isolate after testing positive without symptoms, and 100% would isolate after testing positive if symptomatic. Isolation compliance applied to individuals who were noncompliant with quarantine and to individuals who developed symptoms or tested positive during or after quarantine. The base case assumed a test sensitivity of 70% for symptomatic infections and 60% for asymptomatic infections during infectious phases. We assumed pre-infectious infections were undetectable [2]. The base scenario assumed symptom onset did not occur more than 24h before arrival, but the timing of infection relative to arrival was otherwise random. We assessed 3 additional scenarios regarding the timing of infection relative to arrival and performed extensive scenario analysis (Supplemental Figures S3 – S6 and Tables S1 – S3). Our webtool allows users to customize most parameters based on context-specific data or assumptions. All data and code are public [3].

## RESULTS

A 14-day quarantine with perfect compliance prevented 99.7% (CrI 98.5% – 100.0%) of community transmission risk, but that dropped to 79.7% (78.7 - 80.0%) in the base scenario and 49.8% (49.1 - 50.0%) in a low compliance scenario (Figure). In the base scenario, a 7-day quarantine requirement reduced risk by 71.8% (67.0 - 75.2%) and a 5-day quarantine policy reduced risk by 61.6% (55.6 - 67.0%). The addition of testing generally increased quarantine effectiveness. With perfect compliance, adding a test 24h before quarantine end was most effective, but testing on arrival added more value when compliance was low. Regardless of compliance, the added value of testing declined for longer quarantine durations. With no quarantine, testing on arrival with enforced isolation was most effective, reducing 28.9% (24.7 - 33.7%) of community transmission risk. Testing 72h before arrival reduced risk by only 4.8% (3.5 - 6.1%) in the base scenario and by no more than 10% in any of the scenario analyses. Across all infection timing scenarios, the median proportion of infections that were not yet detectable was over 75% 3 days before arrival and over 39% on arrival, limiting the effectiveness of testing without quarantine (Figure S7).

**Figure.**
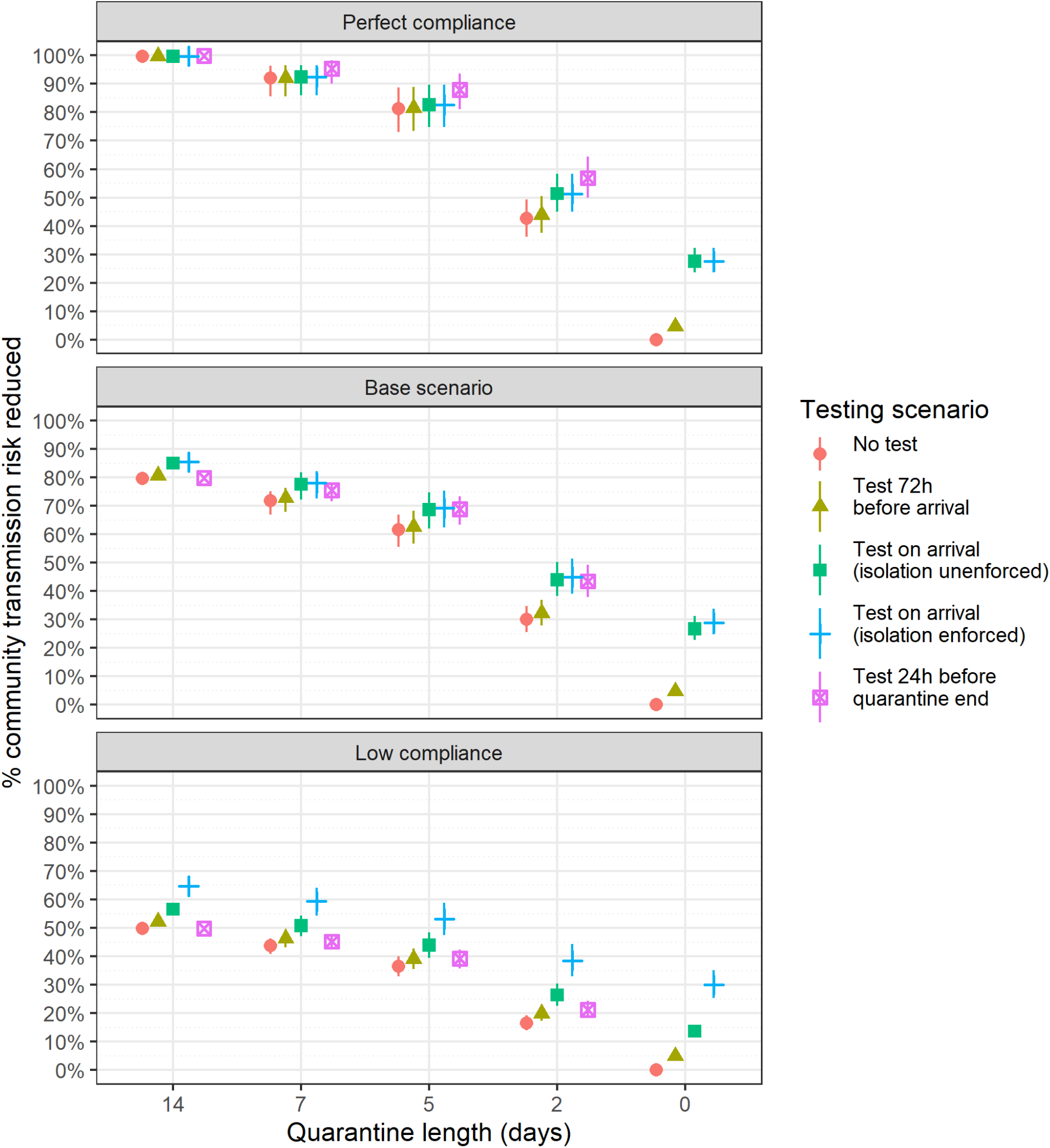
Estimated percent community transmission risk reduction of quarantine and testing policies in a perfect compliance scenario, the base scenario, and a low compliance scenario. The base scenario assumes 80% quarantine compliance; 80% isolation compliance following symptom onset; 90% isolation compliance following a positive test result; and 100% isolation compliance with symptoms and a positive test. The low compliance scenario assumes 50% quarantine compliance; 50% isolation compliance following symptom onset or a positive test result; and 70% isolation compliance following symptom onset and a positive test.

## DISCUSSION

To reduce risk of COVID-19 transmission from arriving travelers, quarantine is more effective than testing alone. Compliance with quarantine and isolation were the most important factors we examined. For example, 5-day quarantine with perfect compliance (81.4%) had a similar median risk reduction to 14-day quarantine in the base case (79.7%). Other model-based analyses recently assessed quarantine and testing policies for arriving travelers and produced similar findings about the ineffectiveness of testing alone [4,5]. Our analysis makes three further contributions. First, our extensive scenario analysis evaluated sensitivity to many unexplored parameters that may be uncertain or context-specific, and our webtool enables policymakers to construct custom scenarios based on local conditions. Second, we analyzed compliance in far greater detail, separately modeling compliance with quarantine and self-isolation following symptom onset or testing positive. Another recent simulation-based analysis found that quarantine and isolation compliance play a similarly critical role in the effectiveness of contact tracing policies [6]. Third, we assessed pre-departure testing to reflect the current US policy of requiring a negative test up to 72 hours before arrival. In all scenarios, most infected travelers were not yet detectable 72h before arriving, limiting the effectiveness of pre-departure testing. However, we did not consider in-transit transmission to other travelers, which pre-departure testing may avert. We also did not consider household transmission during quarantine, a significant risk regardless of quarantine length or testing requirements if arriving travelers cohabitate with community members during quarantine.

By estimating the relative effectiveness of policies to reduce the risk of arriving travelers introducing new SARS-CoV-2 transmission chains, we found that pre-departure testing is minimally effective and compliance with both quarantine and isolation is critical. The optimal policy for a country will depend on characteristics of arriving travelers (e.g., rate of infection, likelihood of importing a new variant) and conditions in the community (e.g., immunization rate, likelihood that imported cases will necessitate economically damaging lockdowns, health system capacity and utilization, and the economic impact of travel restrictions). The webtool we developed allows exploration of the influence of these characteristics in light of local conditions.

## Data Availability

All data and materials have been uploaded to a public repository.

https://doi.org/10.5281/zenodo.4107124

## Appendix

### SUPPLEMENTAL METHODS

#### Calculation of outcomes

This report focuses on the percent community risk reduced by each policy, but the model calculates other outcomes as well:

- **Days at risk of community transmission per infected traveler:** the duration (in days) under each policy for which an infected traveler is in the community (i.e., not in quarantine or isolation) and also in an infectious phase (pre-symptomatic infectious, symptomatic infectious, or asymptomatic infectious)
- **Adjusted days at risk of community transmission per infected traveler:** calculated as above except that asymptomatic infection days in the community are multiplied by the relative risk of transmission for asymptomatic as compared to symptomatic infections.
- **Percent community transmission risk reduced:** The percent decrease in the expectation of the adjusted days at risk of community transmission for a policy as compared to a policy of no quarantine and no testing.

The above outcomes are all independent of the prevalence of infection among travelers and therefore more generalizable. Additional outcomes can be derived for specific scenarios using additional parameters:

- **Days at risk per 10**,**000 arriving travelers:** Estimated by multiplying the days at risk per infected traveler by the prevalence of active infections among travelers, which report per 10,000 travelers in the webtool.
- **Adjusted Days at risk per 10**,**000 arriving travelers:** Estimated by multiplying the adjusted days at risk per infected traveler by the prevalence of active infections among travelers, which report per 10,000 travelers in the webtool.
- **Number of secondary cases per 10**,**000 arriving travelers:** Estimated by multiplying the adjusted days at risk per 10,000 travelers by an estimate for the average number of secondary infections per person-day at risk for a symptomatic infection.

One can derive an estimate of the average number of secondary infections per person-day at risk from an assumed *R*_*t*_, the effective reproductive number for arriving travelers. If travelers’ risk behavior differs from that of local residents, travelers’ *R*_*t*_ may also differ. For instance, in locations allowing indoor dining and tourist activities, travelers may spend considerably more time in indoor public spaces with high transmission risk and therefore may have a higher *R*_*t*_ than the local population. By contrast, it is possible that travelers will have a lower *R*_*t*_ if travelers exhibit fewer high-risk behaviors as compared to the general population (e.g., by disproportionately avoiding public indoor spaces). Our model assumes an average of 5 infectious days per symptomatic individual (.7 in pre-symptomatic infectious days and 3.2 symptomatic infectious days for symptomatic individuals; 5 asymptomatic-infectious days for asymptomatic individuals). Thus, one simple estimate for the number of secondary infections per person-day at risk can then be estimated by dividing the estimated *R*_*t*_ by 5. To factor in the relative risk of transmission for asymptomatic vs. symptomatic infections one could instead factor in the percent asymptomatic infections and the relative transmission risk for asymptomatic vs. symptomatic infectious days.

These additional outcomes are available in the webtool: https://shiny.mchi.mcgill.ca/arussel/quarantineTesting/

#### Relative transmissibility of asymptomatic infections

Researchers in Wanzhou, China, analyzed epidemiologic data for 183 confirmed COVID-19 cases and their close contacts from five generations of transmission, estimating that asymptomatic infections constitute 32.8% of infected persons and cause 19.3% of infections [1]. We calculated the relative risk of transmission for asymptomatic as compared to symptomatic infections as follows:

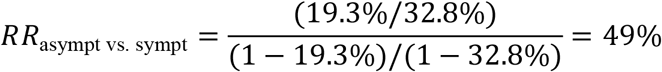

#### Simulation structure

We simulated the infection phases of travelers with asymptomatic infections (non-infectious phase and asymptomatic-infectious phase) and symptomatic infections (non-infectious phase, pre-symptomatic-infectious phase, and symptomatic-infectious phase). We used code from Lauer 2020 to sample 1,000 bootstrapped lognormal parameters for the incubation time distribution [7], which had a mean of 5.1 days. Following Moghadas 2020, we used gamma distributions for the durations of asymptomatic-infectious, pre-symptomatic-infectious, and symptomatic-infectious phases. However, we introduced uncertainty by varying the distributions’ mean and variance uniformly by ±20%, and we sampled 1,000 parameter sets for the duration distributions. The pre-symptomatic-infectious phase had a mean duration of 2.3 days and a variance of 5 days; the symptomatic-infectious phase had a mean duration of 3.2 days and a variance of 3.7 days; and the asymptomatic-infectious phase had a mean duration of 5 days and a variance of 5 days. For the pre-symptomatic-infectious phase, we shifted the gamma distribution by 0.5 days (so the minimum pre-symptomatic-infectious phase duration was 0.5 days) and recalculated shape and scale parameters to match the mean and variance. Figure S1 shows 100 randomly sampled duration distributions.

For each of 1,000 sets of duration distributions, we simulated 5,000 travelers with asymptomatic infection and 10,000 travelers with symptomatic infection. For the symptomatic infections, we sampled from the incubation time distribution to get the time of symptom onset and subtracted the pre-symptomatic-infectious phase to get the time of infectiousness, but we imposed a lower limit that the time of infectiousness was at least 0.5 days after infection. We also sampled the duration of the symptomatic-infectious phase to determine the time each traveler would be no longer infectious. For each traveler with an asymptomatic infection, we determined the time of infectiousness similarly and sampled a duration of asymptomatic infectiousness to determine the time the traveler would no longer be infectious. For each traveler, we then calculated the days in the community and infectious in the pre-symptomatic-infectious, symptomatic-infectious, and asymptomatic-infectious phases using the expressions in the next section. We then took the expected value for each of the 1,000 distribution samples and reported the median and the 1st and 99th percentiles.

#### Calculation of days at risk based on quarantine duration and testing

We calculated the expected number of days at risk of community transmission for each person with an active SARS-CoV-2 infection (either pre-infectious or infectious) at the time of arrival using the expressions below. In these expressions, time is with respect to the time of infection *t*_0_, and we assume infections are not detectable before they are infectious.

##### Asymptomatic infections

We assume travelers with asymptomatic infection comply with quarantine with probability *P*_*q*_. If they do not comply, they will be infectious in the community from the greater of the time they become infectious (*t*_*i*_) and the time they arrive (*u*), and their “infectious days in community” will end when they recover, *t*_*r*_. If they comply with quarantine, their infectious days will begin at the greater of *t*_*i*_ and when quarantine ends, represented by *u* + *d*.

###### Asymptomatic infections with no testing

the expected number of days at risk of community transmission for asymptomatic individuals with no testing is:

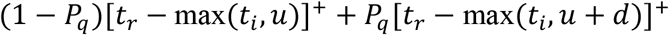

###### Asymptomatic infections with testing 24h before quarantine end

this scenario assumes those who do not comply with quarantine do not receive testing, and those who comply with quarantine are tested 24h before quarantine ends (*t* = *u* + *d* − 1). If they are tested before becoming infectious (*u* + *d* − 1 < *t*_*i*_) they will test negative and be at risk of community transmission from the greater of *t*_*i*_ and *d* + *u* until *t*_*r*_. If they are tested while infectious, they will only be at risk of community transmission if they test false-negative (probability 1 − *Sn*_*A*_ where *Sn*_*A*_ is test sensitivity during the asymptomatic-infectious stage) or if they test positive but refuse to quarantine (*Sn*_*A*_(1 − *P*_*it*_) where *P*_*it*_ is probability of isolation given a positive test and no symptoms). Therefore, the expected number of days at risk of community transmission for asymptomatic individuals with testing 24 hours before end of quarantine is:

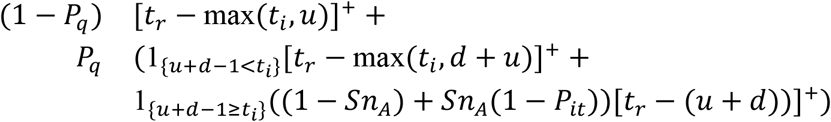

###### Asymptomatic infections with testing on arrival without isolation enforcement

In this scenario, all travelers are tested when they arrive (*t* = *u*). We assume isolation after testing positive is not strictly enforced; as before, the probability of isolating after a positive test is *P*_*it*_. In this testing scenario, the person-days of community risk for a traveler with an asymptomatic infection is:

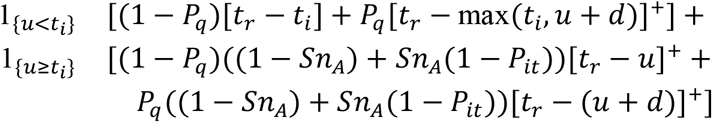

###### Asymptomatic infections with testing on arrival with isolation enforcement

In this scenario, all travelers are tested when they arrive (*t* = *u*). We assume those who test positive on arrival are either denied entry or have quarantine strictly enforced. The person-days of community risk for a traveler with an asymptomatic infection is:

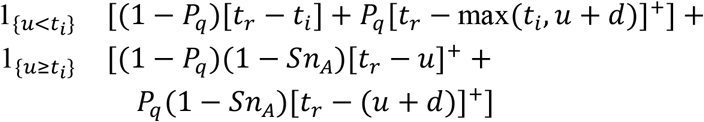

###### Asymptomatic infections with testing 72h before arrival

In this scenario, travelers are tested 3 days before arrival (*t* = *u* − 3). We assume those who test positive will be denied entry.

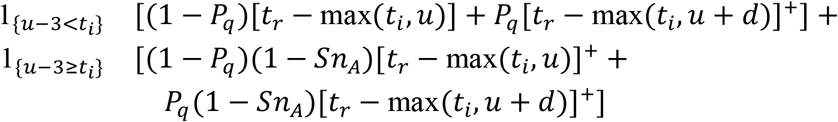

##### Symptomatic infections

For travelers with symptomatic infections, we calculate the days at risk of community transmission separately for the pre-symptomatic and symptomatic infection phases. Days at risk during the pre-symptomatic stage are calculated the same as asymptomatic days except that the endpoint for pre-symptomatic days at risk is the time that symptoms begin (*t*_*s*_) rather than the time of recovery (*t*_*r*_), and we use test sensitivity for detecting an infection in the pre-symptomatic-infectious stage (*Sn*_*P*_). In calculating symptomatic days in the community, we consider that travelers are required to isolate once symptoms develop, and we factor in the probability of compliance with isolation after symptom onset *P*_*is*_.

###### Symptomatic infections with no testing

the expected number of days at risk of community transmission while in **pre-symptomatic phase** with no testing is:

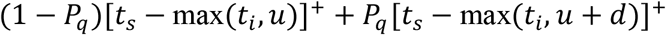

We assume travelers will isolate while in symptomatic-infectious stage with probability *P*_*is*_. Days at risk while symptomatic begin at *t*_*s*_, the time symptoms begin, and end at *t*_*r*_, the time of recovery when individuals are no longer infectious. Therefore, the expected number of days at risk of community transmission while in **symptomatic phase** with no testing is:

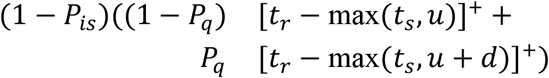

###### Symptomatic infections with testing 24h before end of quarantine

With testing 24h before quarantine end, we assume that the days at risk for those who do not comply with quarantine (probability (1 − *P*_*q*_)) is unchanged because they also do not get tested. Those who comply will test negative if they are tested before infectious (*u* + *d* − 1 < *t*_*i*_). With probability 1 − *P*_*is*_ they will not isolate after developing symptoms and will incur days at risk until *t*_*r*_. If tested during pre-symptomatic infectious stage (*t*_*i*_ ≤ *u* + *d* − 1 < *t*_*s*_), they will incur days at risk while symptomatic if they test false negative (probability (1 − *Sn*_*P*_)) and they do not isolate (probability (1 − *P*_*is*_) or if they test positive and refuse to isolate (*Sn*_*P*_(1 − *P*_*ib*_), where *P*_*ib*_ is probability of isolating with both a positive test and symptoms). For those tested while symptomatic, they will incur days at risk if the test is false negative (1 − *Sn*_*S*_ where *Sn*_*S*_ is test sensitivity for those in symptomatic-infectious state) or if they test positive and refuse to isolate (*Sn*_*S*_(1 − *P*_*ib*_)). with testing 24h before the end of quarantine, the expected number of days at risk of community transmission while in **pre-symptomatic phase** is:

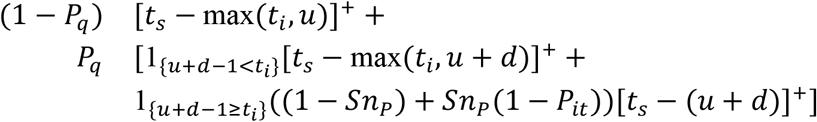

The expected number of days at risk of community transmission while in the **symptomatic-infectious phase** with testing 24 hours before end of quarantine is:

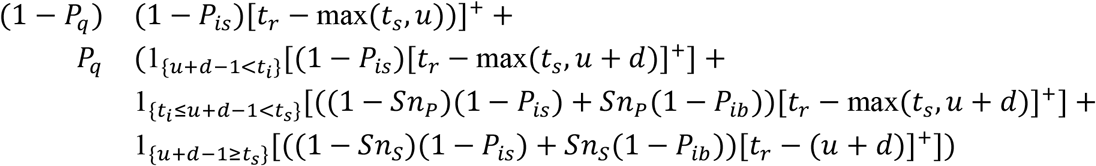

###### Symptomatic infections with testing on arrival without isolation enforcement

the expected number of days at risk of community transmission while in the **pre-symptomatic phase** is:

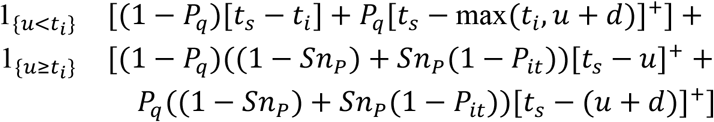

The expected number of days at risk of community transmission while in the **symptomatic phase** is:

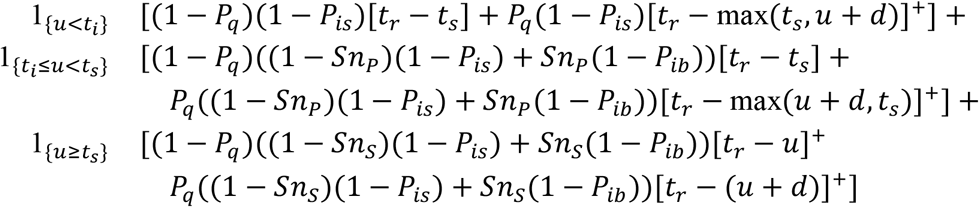

###### Symptomatic infections with testing on arrival with isolation enforcement

the expected number of days at risk of community transmission while in the **pre-symptomatic phase** is:

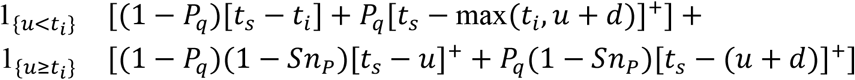

The expected number of days at risk of community transmission while in the **symptomatic phase** is:

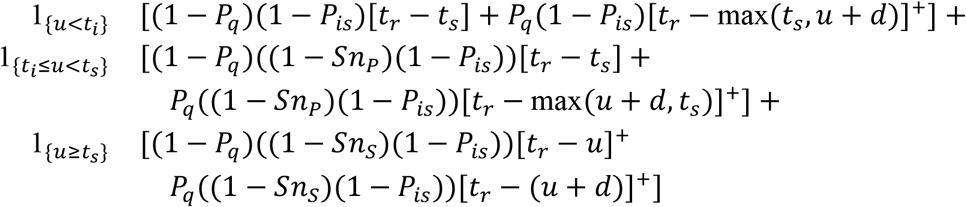

###### Symptomatic infections with testing 72h before arrival

the expected number of days at risk of community transmission while in the **pre-symptomatic phase** is:

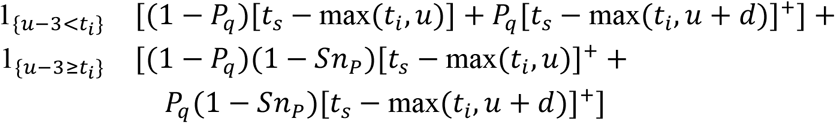

The expected number of days at risk of community transmission while in the **symptomatic phase** is:

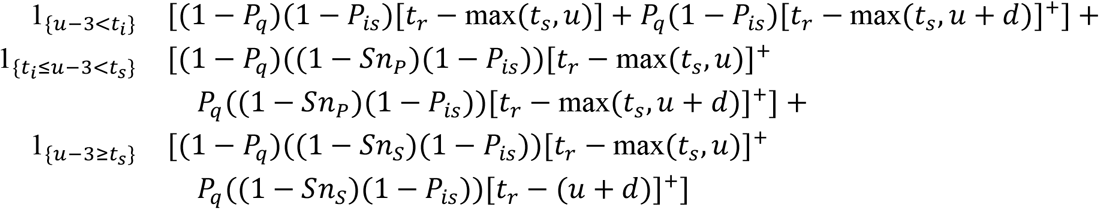

## SUPPLEMENTAL TABLES

**Table S1.** Estimated percent of transmission risk reduced for quarantine and testing policies in 11 modeling scenarios. Median and 98% credible interval provided. Abbreviation: UE, unenforced.

| Test policy | 0 days | 2 days | 5 days | 7 days | 14 days |
| --- | --- | --- | --- | --- | --- |
| 10% of infections are asymptomatic |  |  |  |  |  |
| None | 0.0% (0.0 - 0.0%) | 31.1% (26.6 - 36.2%) | 63.1% (57.1 - 69.0%) | 72.7% (67.9 - 76.2%) | 79.8% (78.8 - 80.0%) |
| 3 days prior | 3.6% (2.7 - 4.6%) | 32.9% (28.3 - 37.9%) | 64.0% (58.1 - 69.9%) | 73.5% (68.8 - 76.9%) | 80.5% (79.5 - 80.8%) |
| On arrival UE | 27.1% (23.0 - 31.6%) | 45.1% (39.1 - 51.6%) | 70.3% (63.6 - 76.7%) | 78.7% (73.5 - 82.8%) | 85.1% (83.6 - 86.2%) |
| On arrival enforced | 28.9% (24.6 - 33.7%) | 45.9% (39.7 - 52.5%) | 70.7% (63.9 - 77.1%) | 79.1% (73.8 - 83.2%) | 85.5% (83.9 - 86.7%) |
| At end |  | 44.7% (39.0 - 50.8%) | 70.2% (64.8 - 74.8%) | 76.3% (72.4 - 78.5%) | 79.9% (79.2 - 80.0%) |
| 100% test sensitivity during infectious disease phases (pre-infectious infections not detectable) |  |  |  |  |  |
| None | 0.0% (0.0 - 0.0%) | 30.0% (25.6 - 34.9%) | 61.6% (55.6 - 67.0%) | 71.8% (67.0 - 75.2%) | 79.7% (78.7 - 80.0%) |
| 3 days prior | 7.5% (5.5 - 9.7%) | 33.5% (29.1 - 38.2%) | 63.3% (57.4 - 68.9%) | 73.3% (68.5 - 76.8%) | 81.2% (80.1 - 81.8%) |
| On arrival UE | 40.5% (34.8 - 47.3%) | 51.3% (44.7 - 58.4%) | 72.3% (65.4 - 78.9%) | 80.6% (75.0 - 85.2%) | 87.8% (85.9 - 89.3%) |
| On arrival enforced | 43.8% (37.7 - 51.2%) | 52.9% (46.0 - 60.3%) | 73.1% (66.1 - 79.9%) | 81.4% (75.6 - 86.1%) | 88.4% (86.5 - 90.1%) |
| At end |  | 50.5% (44.2 - 57.0%) | 72.8% (67.6 - 77.0%) | 77.6% (73.9 - 79.5%) | 79.9% (79.4 - 80.0%) |
| 70% of infections are asymptomatic |  |  |  |  |  |
| None | 0.0% (0.0 - 0.0%) | 28.9% (24.5 - 33.7%) | 60.0% (53.8 - 65.7%) | 70.8% (65.7 - 74.7%) | 79.7% (78.5 - 79.9%) |
| 3 days prior | 5.9% (4.2 - 8.0%) | 31.6% (27.4 - 36.2%) | 61.4% (55.2 - 67.0%) | 72.1% (67.1 - 75.7%) | 80.8% (79.7 - 81.4%) |
| On arrival UE | 26.3% (22.5 - 30.9%) | 42.8% (37.3 - 49.0%) | 66.9% (60.4 - 73.2%) | 76.5% (71.2 - 80.9%) | 84.9% (83.3 - 86.0%) |
| On arrival enforced | 28.8% (24.6 - 33.8%) | 44.1% (38.5 - 50.4%) | 67.6% (61.0 - 73.8%) | 77.1% (71.7 - 81.5%) | 85.4% (83.8 - 86.6%) |
| At end |  | 42.2% (36.9 - 47.9%) | 67.7% (62.0 - 72.4%) | 74.8% (70.6 - 77.5%) | 79.8% (79.0 - 80.0%) |
| All infections are asymptomatic |  |  |  |  |  |
| None | 0.0% (0.0 - 0.0%) | 27.7% (23.4 - 32.6%) | 58.4% (52.0 - 64.5%) | 69.8% (64.4 - 74.2%) | 79.6% (78.4 - 80.0%) |
| 3 days prior | 7.1% (4.7 - 9.9%) | 31.0% (26.6 - 35.7%) | 60.1% (53.9 - 66.2%) | 71.3% (66.1 - 75.5%) | 81.0% (79.7 - 81.8%) |
| On arrival UE | 25.8% (21.7 - 30.6%) | 41.6% (36.3 - 47.3%) | 65.2% (58.6 - 71.6%) | 75.4% (69.7 - 80.1%) | 84.7% (83.1 - 86.0%) |
| On arrival enforced | 28.7% (24.1 - 34.0%) | 43.2% (37.7 - 49.0%) | 65.9% (59.3 - 72.3%) | 76.0% (70.3 - 80.8%) | 85.3% (83.6 - 86.7%) |
| At end |  | 40.9% (35.8 - 46.3%) | 66.3% (60.9 - 71.2%) | 74.1% (69.7 - 77.1%) | 79.8% (78.9 - 80.0%) |
| All infections are symptomatic |  |  |  |  |  |
| None | 0.0% (0.0 - 0.0%) | 31.5% (26.7 - 36.8%) | 63.6% (57.3 - 69.4%) | 73.0% (68.3 - 76.4%) | 79.8% (78.9 - 80.0%) |
| 3 days prior | 3.2% (2.3 - 4.3%) | 33.1% (28.5 - 38.2%) | 64.4% (58.3 - 70.3%) | 73.7% (69.1 - 77.1%) | 80.4% (79.5 - 80.7%) |
| On arrival UE | 27.2% (23.0 - 31.7%) | 45.5% (39.4 - 52.1%) | 70.9% (64.2 - 77.3%) | 79.0% (73.9 - 83.1%) | 85.2% (83.6 - 86.3%) |
| On arrival enforced | 29.0% (24.5 - 34.0%) | 46.1% (39.9 - 52.9%) | 71.2% (64.6 - 77.7%) | 79.4% (74.2 - 83.5%) | 85.5% (83.9 - 86.7%) |
| At end |  | 45.1% (39.4 - 51.3%) | 70.6% (65.1 - 75.3%) | 76.5% (72.6 - 78.7%) | 79.9% (79.3 - 80.0%) |
| Base scenario |  |  |  |  |  |
| None | 0.0% (0.0 - 0.0%) | 30.0% (25.6 - 34.9%) | 61.6% (55.6 - 67.0%) | 71.8% (67.0 - 75.2%) | 79.7% (78.7 - 80.0%) |
| 3 days prior | 4.8% (3.5 - 6.1%) | 32.2% (27.9 - 36.9%) | 62.7% (56.7 - 68.2%) | 72.8% (68.0 - 76.2%) | 80.7% (79.6 - 81.1%) |
| On arrival UE | 26.7% (22.9 - 31.2%) | 44.0% (38.3 - 50.3%) | 68.7% (62.0 - 74.8%) | 77.6% (72.2 - 81.7%) | 85.0% (83.5 - 86.1%) |
| On arrival enforced | 28.9% (24.7 - 33.7%) | 45.0% (39.1 - 51.4%) | 69.2% (62.5 - 75.4%) | 78.1% (72.6 - 82.3%) | 85.4% (83.8 - 86.6%) |
| At end |  | 43.5% (37.9 - 49.3%) | 68.9% (63.4 - 73.4%) | 75.5% (71.5 - 77.9%) | 79.9% (79.1 - 80.0%) |
| Infected at time of arrival |  |  |  |  |  |
| None | 0.0% (0.0 - 0.0%) | 8.2% (5.5 - 11.8%) | 38.2% (29.1 - 48.2%) | 56.2% (46.2 - 65.3%) | 78.5% (74.7 - 79.7%) |
| 3 days prior | 0.0% (0.0 - 0.0%) | 8.2% (5.5 - 11.8%) | 38.2% (29.1 - 48.2%) | 56.2% (46.2 - 65.3%) | 78.5% (74.7 - 79.7%) |
| On arrival UE | 0.0% (0.0 - 0.0%) | 8.2% (5.5 - 11.8%) | 38.2% (29.1 - 48.2%) | 56.2% (46.2 - 65.3%) | 78.5% (74.7 - 79.7%) |
| On arrival enforced | 0.0% (0.0 - 0.0%) | 8.2% (5.5 - 11.8%) | 38.2% (29.1 - 48.2%) | 56.2% (46.2 - 65.3%) | 78.5% (74.7 - 79.7%) |
| At end |  | 16.7% (11.8 - 23.0%) | 49.0% (38.9 - 59.4%) | 64.6% (55.3 - 72.4%) | 79.2% (76.5 - 79.9%) |
| Low compliance (50% for quarantine, with symptoms, or with a positive test; 70% for both symptoms and a positive test) |  |  |  |  |  |
| None | 0.0% (0.0 - 0.0%) | 16.5% (14.0 - 19.2%) | 36.5% (32.9 - 40.0%) | 43.8% (40.8 - 46.3%) | 49.8% (49.1 - 50.0%) |
| 3 days prior | 4.9% (3.6 - 6.3%) | 20.0% (17.2 - 23.0%) | 39.2% (35.4 - 42.7%) | 46.3% (43.2 - 49.0%) | 52.2% (51.2 - 53.0%) |
| On arrival UE | 13.6% (11.5 - 16.0%) | 26.4% (22.6 - 30.4%) | 44.0% (39.4 - 48.4%) | 50.8% (47.1 - 54.3%) | 56.5% (55.1 - 57.9%) |
| On arrival enforced | 30.0% (25.4 - 35.1%) | 38.5% (32.9 - 44.3%) | 53.1% (47.4 - 58.9%) | 59.3% (54.3 - 64.1%) | 64.7% (62.1 - 67.4%) |
| At end |  | 21.2% (18.3 - 24.2%) | 39.2% (35.8 - 42.4%) | 45.2% (42.5 - 47.2%) | 49.8% (49.3 - 50.0%) |
| Perfect compliance (100% compliance for quarantine and isolation) |  |  |  |  |  |
| None | 0.0% (0.0 - 0.0%) | 42.7% (36.2 - 49.4%) | 81.4% (73.1 - 88.7%) | 92.1% (85.6 - 96.4%) | 99.7% (98.5 - 100.0%) |
| 3 days prior | 4.6% (3.4 - 6.2%) | 43.9% (37.6 - 50.4%) | 81.5% (73.3 - 88.8%) | 92.1% (85.7 - 96.5%) | 99.7% (98.5 - 100.0%) |
| On arrival UE | 27.6% (23.7 - 32.3%) | 51.4% (45.0 - 58.4%) | 82.6% (74.7 - 89.7%) | 92.4% (86.0 - 96.6%) | 99.7% (98.5 - 100.0%) |
| On arrival enforced | 27.6% (23.7 - 32.3%) | 51.4% (45.0 - 58.4%) | 82.6% (74.7 - 89.7%) | 92.4% (86.0 - 96.6%) | 99.7% (98.5 - 100.0%) |
| At end |  | 56.9% (49.9 - 64.5%) | 87.9% (81.0 - 93.6%) | 95.3% (90.1 - 98.3%) | 99.9% (99.0 - 100.0%) |
| Travelers are equally likely to be in any disease phase (including symptomatic) |  |  |  |  |  |
| None | 0.0% (0.0 - 0.0%) | 30.9% (26.4 - 35.8%) | 62.0% (55.8 - 67.6%) | 72.1% (66.9 - 75.6%) | 79.7% (78.6 - 80.0%) |
| 3 days prior | 6.6% (5.1 - 8.3%) | 33.8% (29.3 - 38.7%) | 63.6% (57.3 - 69.2%) | 73.4% (68.4 - 76.9%) | 81.0% (79.9 - 81.5%) |
| On arrival UE | 27.6% (23.9 - 32.0%) | 44.9% (39.2 - 51.1%) | 69.1% (62.5 - 75.2%) | 77.9% (72.4 - 82.1%) | 85.2% (83.5 - 86.3%) |
| On arrival enforced | 29.8% (25.9 - 34.6%) | 45.9% (40.1 - 52.1%) | 69.6% (62.9 - 75.9%) | 78.4% (72.8 - 82.6%) | 85.6% (84.0 - 86.8%) |
| At end |  | 43.8% (38.3 - 49.6%) | 68.8% (63.2 - 73.4%) | 75.5% (71.3 - 78.0%) | 79.9% (79.1 - 80.0%) |
| Travelers have not developed symptoms before arriving |  |  |  |  |  |
| None | 0.0% (0.0 - 0.0%) | 30.1% (25.5 - 34.8%) | 61.7% (55.6 - 67.2%) | 71.9% (67.1 - 75.3%) | 79.7% (78.7 - 80.0%) |
| 3 days prior | 4.1% (3.0 - 5.3%) | 31.9% (27.5 - 36.6%) | 62.7% (56.6 - 68.1%) | 72.7% (68.1 - 76.1%) | 80.5% (79.6 - 80.9%) |
| On arrival UE | 25.4% (21.4 - 30.0%) | 43.2% (37.3 - 49.6%) | 68.5% (61.7 - 74.7%) | 77.5% (72.1 - 81.6%) | 84.8% (83.2 - 85.9%) |
| On arrival enforced | 27.7% (23.3 - 32.6%) | 44.2% (38.1 - 50.8%) | 69.0% (62.1 - 75.3%) | 77.9% (72.5 - 82.1%) | 85.2% (83.6 - 86.4%) |
| At end |  | 43.2% (37.7 - 49.2%) | 69.1% (63.6 - 73.5%) | 75.6% (71.8 - 78.0%) | 79.9% (79.2 - 80.0%) |

**Table S2.** Expected days at risk of community transmission risk per infectious traveler for quarantine and testing policies in 11 modeling scenarios. Median and 98% credible interval provided. Abbreviation: UE, unenforced.

| Test policy | 0 days | 2 days | 5 days | 7 days | 14 days |
| --- | --- | --- | --- | --- | --- |
| 10% of infections are asymptomatic |  |  |  |  |  |
| None | 2.0 (1.7 - 2.3) | 1.4 (1.1 - 1.6) | 0.73 (0.58 - 0.94) | 0.54 (0.43 - 0.69) | 0.40 (0.34 - 0.47) |
| 3 days prior | 1.9 (1.6 - 2.2) | 1.3 (1.1 - 1.6) | 0.71 (0.56 - 0.91) | 0.52 (0.42 - 0.67) | 0.38 (0.33 - 0.45) |
| On arrival UE | 1.4 (1.2 - 1.6) | 1.1 (0.90 - 1.3) | 0.59 (0.45 - 0.77) | 0.42 (0.33 - 0.57) | 0.29 (0.25 - 0.35) |
| On arrival enforced | 1.4 (1.2 - 1.6) | 1.1 (0.88 - 1.3) | 0.58 (0.44 - 0.76) | 0.41 (0.32 - 0.56) | 0.28 (0.24 - 0.34) |
| At end |  | 1.1 (0.91 - 1.3) | 0.59 (0.47 - 0.76) | 0.47 (0.38 - 0.60) | 0.40 (0.34 - 0.46) |
| 100% test sensitivity during infectious disease phases (pre-infectious infections not detectable) |  |  |  |  |  |
| None | 2.4 (2.1 - 2.8) | 1.7 (1.4 - 2.0) | 0.96 (0.75 - 1.2) | 0.70 (0.56 - 0.90) | 0.50 (0.43 - 0.58) |
| 3 days prior | 2.2 (1.9 - 2.5) | 1.6 (1.4 - 1.9) | 0.90 (0.70 - 1.2) | 0.65 (0.52 - 0.85) | 0.45 (0.40 - 0.52) |
| On arrival UE | 1.4 (1.2 - 1.7) | 1.2 (0.97 - 1.4) | 0.69 (0.51 - 0.93) | 0.48 (0.35 - 0.67) | 0.29 (0.25 - 0.36) |
| On arrival enforced | 1.3 (1.1 - 1.6) | 1.1 (0.93 - 1.4) | 0.67 (0.49 - 0.91) | 0.46 (0.33 - 0.65) | 0.27 (0.23 - 0.34) |
| At end |  | 1.2 (1.00 - 1.5) | 0.67 (0.53 - 0.87) | 0.55 (0.46 - 0.70) | 0.49 (0.42 - 0.56) |
| 70% of infections are asymptomatic |  |  |  |  |  |
| None | 2.9 (2.5 - 3.4) | 2.1 (1.7 - 2.5) | 1.2 (0.88 - 1.6) | 0.86 (0.65 - 1.1) | 0.59 (0.50 - 0.71) |
| 3 days prior | 2.7 (2.3 - 3.1) | 2.0 (1.6 - 2.4) | 1.1 (0.85 - 1.5) | 0.82 (0.62 - 1.1) | 0.55 (0.47 - 0.66) |
| On arrival UE | 2.1 (1.8 - 2.5) | 1.7 (1.3 - 2.1) | 0.97 (0.71 - 1.3) | 0.69 (0.51 - 0.97) | 0.44 (0.37 - 0.54) |
| On arrival enforced | 2.1 (1.8 - 2.4) | 1.6 (1.3 - 2.0) | 0.95 (0.69 - 1.3) | 0.67 (0.49 - 0.95) | 0.42 (0.35 - 0.52) |
| At end |  | 1.7 (1.4 - 2.1) | 0.95 (0.72 - 1.3) | 0.74 (0.58 - 0.97) | 0.59 (0.49 - 0.69) |
| All infections are asymptomatic |  |  |  |  |  |
| None | 3.4 (2.7 - 4.0) | 2.4 (1.9 - 3.1) | 1.4 (1.00 - 1.9) | 1.0 (0.73 - 1.4) | 0.69 (0.56 - 0.85) |
| 3 days prior | 3.1 (2.6 - 3.7) | 2.3 (1.8 - 2.9) | 1.3 (0.96 - 1.9) | 0.96 (0.69 - 1.4) | 0.64 (0.52 - 0.78) |
| On arrival UE | 2.5 (2.0 - 3.0) | 2.0 (1.5 - 2.5) | 1.2 (0.82 - 1.7) | 0.82 (0.57 - 1.2) | 0.51 (0.41 - 0.65) |
| On arrival enforced | 2.4 (2.0 - 2.9) | 1.9 (1.5 - 2.4) | 1.1 (0.79 - 1.6) | 0.80 (0.56 - 1.2) | 0.49 (0.40 - 0.63) |
| At end |  | 2.0 (1.5 - 2.5) | 1.1 (0.82 - 1.5) | 0.87 (0.65 - 1.2) | 0.68 (0.55 - 0.83) |
| All infections are symptomatic |  |  |  |  |  |
| None | 1.8 (1.5 - 2.2) | 1.2 (0.99 - 1.5) | 0.66 (0.49 - 0.87) | 0.49 (0.37 - 0.64) | 0.37 (0.30 - 0.44) |
| 3 days prior | 1.7 (1.5 - 2.1) | 1.2 (0.97 - 1.5) | 0.64 (0.48 - 0.85) | 0.47 (0.36 - 0.63) | 0.35 (0.29 - 0.43) |
| On arrival UE | 1.3 (1.1 - 1.5) | 0.98 (0.79 - 1.2) | 0.52 (0.38 - 0.71) | 0.38 (0.28 - 0.52) | 0.27 (0.22 - 0.32) |
| On arrival enforced | 1.3 (1.1 - 1.5) | 0.97 (0.78 - 1.2) | 0.52 (0.38 - 0.70) | 0.37 (0.28 - 0.51) | 0.26 (0.22 - 0.32) |
| At end |  | 0.99 (0.80 - 1.2) | 0.53 (0.40 - 0.70) | 0.43 (0.33 - 0.55) | 0.36 (0.30 - 0.44) |
| Base scenario |  |  |  |  |  |
| None | 2.4 (2.1 - 2.8) | 1.7 (1.4 - 2.0) | 0.96 (0.75 - 1.2) | 0.70 (0.56 - 0.90) | 0.50 (0.43 - 0.58) |
| 3 days prior | 2.3 (2.0 - 2.6) | 1.7 (1.4 - 1.9) | 0.92 (0.72 - 1.2) | 0.67 (0.53 - 0.87) | 0.47 (0.41 - 0.55) |
| On arrival UE | 1.8 (1.6 - 2.0) | 1.4 (1.1 - 1.6) | 0.78 (0.59 - 1.0) | 0.56 (0.42 - 0.76) | 0.37 (0.32 - 0.44) |
| On arrival enforced | 1.7 (1.5 - 2.0) | 1.3 (1.1 - 1.6) | 0.77 (0.58 - 1.0) | 0.55 (0.41 - 0.74) | 0.35 (0.31 - 0.43) |
| At end |  | 1.4 (1.2 - 1.7) | 0.77 (0.61 - 0.99) | 0.60 (0.50 - 0.77) | 0.49 (0.42 - 0.57) |
| Infected at time of arrival |  |  |  |  |  |
| None | 3.6 (3.0 - 4.1) | 3.3 (2.7 - 3.8) | 2.3 (1.8 - 2.8) | 1.6 (1.2 - 2.1) | 0.78 (0.63 - 0.99) |
| 3 days prior | 3.6 (3.0 - 4.1) | 3.3 (2.7 - 3.8) | 2.3 (1.8 - 2.8) | 1.6 (1.2 - 2.1) | 0.78 (0.63 - 0.99) |
| On arrival UE | 3.6 (3.0 - 4.1) | 3.3 (2.7 - 3.8) | 2.3 (1.8 - 2.8) | 1.6 (1.2 - 2.1) | 0.78 (0.63 - 0.99) |
| On arrival enforced | 3.6 (3.0 - 4.1) | 3.3 (2.7 - 3.8) | 2.3 (1.8 - 2.8) | 1.6 (1.2 - 2.1) | 0.78 (0.63 - 0.99) |
| At end |  | 3.0 (2.5 - 3.5) | 1.9 (1.5 - 2.4) | 1.3 (0.97 - 1.8) | 0.75 (0.61 - 0.92) |
| Low compliance (50% for quarantine, with symptoms, or with a positive test; 70% for both symptoms and a positive test) |  |  |  |  |  |
| None | 3.0 (2.6 - 3.4) | 2.5 (2.1 - 2.9) | 1.9 (1.6 - 2.2) | 1.7 (1.4 - 2.0) | 1.5 (1.3 - 1.7) |
| 3 days prior | 2.8 (2.5 - 3.2) | 2.4 (2.0 - 2.7) | 1.8 (1.5 - 2.1) | 1.6 (1.4 - 1.9) | 1.4 (1.2 - 1.6) |
| On arrival UE | 2.6 (2.2 - 2.9) | 2.2 (1.9 - 2.6) | 1.7 (1.4 - 2.0) | 1.5 (1.2 - 1.7) | 1.3 (1.1 - 1.5) |
| On arrival enforced | 2.1 (1.8 - 2.4) | 1.8 (1.6 - 2.2) | 1.4 (1.2 - 1.7) | 1.2 (1.0 - 1.5) | 1.1 (0.92 - 1.2) |
| At end |  | 2.4 (2.0 - 2.7) | 1.8 (1.5 - 2.1) | 1.6 (1.4 - 1.9) | 1.5 (1.3 - 1.7) |
| Perfect compliance (100% compliance for quarantine and isolation) |  |  |  |  |  |
| None | 2.1 (1.7 - 2.4) | 1.2 (0.98 - 1.5) | 0.44 (0.26 - 0.67) | 0.19 (0.090 - 0.36) | 0.007 (0.001 - 0.036) |
| 3 days prior | 1.9 (1.6 - 2.2) | 1.2 (0.96 - 1.5) | 0.43 (0.26 - 0.67) | 0.19 (0.089 - 0.36) | 0.007 (0.001 - 0.036) |
| On arrival UE | 1.5 (1.3 - 1.7) | 1.0 (0.82 - 1.3) | 0.40 (0.24 - 0.63) | 0.19 (0.085 - 0.35) | 0.007 (0.001 - 0.036) |
| On arrival enforced | 1.5 (1.3 - 1.7) | 1.0 (0.82 - 1.3) | 0.40 (0.24 - 0.63) | 0.19 (0.085 - 0.35) | 0.007 (0.001 - 0.036) |
| At end |  | 0.90 (0.71 - 1.1) | 0.27 (0.15 - 0.45) | 0.11 (0.043 - 0.23) | 0.003 (0.000 - 0.024) |
| Travelers are equally likely to be in any disease phase (including symptomatic) |  |  |  |  |  |
| None | 2.2 (1.9 - 2.5) | 1.5 (1.3 - 1.8) | 0.86 (0.67 - 1.1) | 0.63 (0.49 - 0.82) | 0.45 (0.38 - 0.53) |
| 3 days prior | 2.0 (1.8 - 2.3) | 1.5 (1.2 - 1.7) | 0.82 (0.64 - 1.1) | 0.60 (0.47 - 0.79) | 0.42 (0.36 - 0.49) |
| On arrival UE | 1.6 (1.4 - 1.8) | 1.2 (1.0 - 1.5) | 0.70 (0.52 - 0.94) | 0.50 (0.37 - 0.69) | 0.33 (0.28 - 0.39) |
| On arrival enforced | 1.5 (1.3 - 1.8) | 1.2 (0.99 - 1.5) | 0.69 (0.51 - 0.93) | 0.49 (0.36 - 0.67) | 0.32 (0.27 - 0.38) |
| At end |  | 1.2 (1.0 - 1.5) | 0.70 (0.55 - 0.91) | 0.55 (0.44 - 0.70) | 0.44 (0.38 - 0.52) |
| Travelers have not developed symptoms before arriving |  |  |  |  |  |
| None | 2.6 (2.2 - 2.9) | 1.8 (1.5 - 2.1) | 1.0 (0.80 - 1.3) | 0.74 (0.60 - 0.94) | 0.52 (0.45 - 0.61) |
| 3 days prior | 2.5 (2.1 - 2.8) | 1.8 (1.5 - 2.0) | 0.98 (0.77 - 1.2) | 0.71 (0.57 - 0.91) | 0.50 (0.43 - 0.58) |
| On arrival UE | 1.9 (1.7 - 2.2) | 1.5 (1.2 - 1.7) | 0.83 (0.64 - 1.1) | 0.59 (0.46 - 0.79) | 0.39 (0.34 - 0.46) |
| On arrival enforced | 1.8 (1.6 - 2.1) | 1.4 (1.2 - 1.7) | 0.82 (0.63 - 1.1) | 0.58 (0.44 - 0.78) | 0.38 (0.33 - 0.45) |
| At end |  | 1.5 (1.2 - 1.7) | 0.81 (0.65 - 1.0) | 0.64 (0.52 - 0.81) | 0.52 (0.45 - 0.60) |

**Table S3.** Expected adjusted days at risk of community transmission risk per infectious traveler for quarantine and testing policies in 11 modeling scenarios. Days at risk for travelers with asymptomatic infections are multiplied by the relative transmission risk of asymptomatic as compared to symptomatic infections (49% in base case). Median and 98% credible interval provided. Abbreviation: UE, unenforced.

| Test policy | 0 days | 2 days | 5 days | 7 days | 14 days |
| --- | --- | --- | --- | --- | --- |
| 10% of infections are asymptomatic |  |  |  |  |  |
| None | 1.8 (1.5 - 2.1) | 1.2 (1.0 - 1.5) | 0.66 (0.51 - 0.86) | 0.49 (0.38 - 0.64) | 0.36 (0.30 - 0.43) |
| 3 days prior | 1.7 (1.5 - 2.0) | 1.2 (0.98 - 1.4) | 0.64 (0.50 - 0.83) | 0.48 (0.37 - 0.62) | 0.35 (0.29 - 0.42) |
| On arrival UE | 1.3 (1.1 - 1.5) | 0.98 (0.80 - 1.2) | 0.53 (0.40 - 0.70) | 0.38 (0.29 - 0.51) | 0.27 (0.23 - 0.32) |
| On arrival enforced | 1.3 (1.1 - 1.5) | 0.96 (0.79 - 1.2) | 0.52 (0.39 - 0.69) | 0.37 (0.28 - 0.51) | 0.26 (0.22 - 0.31) |
| At end |  | 0.99 (0.81 - 1.2) | 0.53 (0.41 - 0.70) | 0.43 (0.34 - 0.55) | 0.36 (0.30 - 0.43) |
| 100% test sensitivity during infectious disease phases (pre-infectious infections not detectable) |  |  |  |  |  |
| None | 1.7 (1.5 - 2.0) | 1.2 (1.0 - 1.4) | 0.67 (0.54 - 0.84) | 0.49 (0.40 - 0.63) | 0.35 (0.31 - 0.41) |
| 3 days prior | 1.6 (1.4 - 1.8) | 1.2 (0.98 - 1.4) | 0.64 (0.51 - 0.80) | 0.46 (0.38 - 0.60) | 0.33 (0.29 - 0.38) |
| On arrival UE | 1.0 (0.90 - 1.2) | 0.84 (0.70 - 1.0) | 0.48 (0.36 - 0.65) | 0.34 (0.25 - 0.47) | 0.21 (0.18 - 0.26) |
| On arrival enforced | 0.97 (0.84 - 1.1) | 0.82 (0.68 - 0.99) | 0.47 (0.34 - 0.63) | 0.32 (0.24 - 0.46) | 0.20 (0.17 - 0.25) |
| At end |  | 0.86 (0.72 - 1.0) | 0.47 (0.37 - 0.62) | 0.39 (0.33 - 0.50) | 0.35 (0.30 - 0.41) |
| 70% of infections are asymptomatic |  |  |  |  |  |
| None | 1.7 (1.5 - 1.9) | 1.2 (1.00 - 1.4) | 0.67 (0.52 - 0.88) | 0.49 (0.39 - 0.65) | 0.35 (0.30 - 0.41) |
| 3 days prior | 1.6 (1.4 - 1.8) | 1.2 (0.96 - 1.4) | 0.65 (0.50 - 0.84) | 0.47 (0.37 - 0.62) | 0.33 (0.28 - 0.38) |
| On arrival UE | 1.2 (1.1 - 1.4) | 0.96 (0.80 - 1.2) | 0.56 (0.42 - 0.74) | 0.40 (0.30 - 0.55) | 0.26 (0.22 - 0.31) |
| On arrival enforced | 1.2 (1.0 - 1.4) | 0.94 (0.78 - 1.1) | 0.55 (0.41 - 0.73) | 0.39 (0.29 - 0.54) | 0.25 (0.21 - 0.30) |
| At end |  | 0.97 (0.81 - 1.2) | 0.55 (0.43 - 0.71) | 0.43 (0.34 - 0.55) | 0.34 (0.29 - 0.40) |
| All infections are asymptomatic |  |  |  |  |  |
| None | 1.7 (1.3 - 2.0) | 1.2 (0.92 - 1.5) | 0.68 (0.49 - 0.94) | 0.50 (0.36 - 0.69) | 0.34 (0.27 - 0.42) |
| 3 days prior | 1.5 (1.3 - 1.8) | 1.1 (0.88 - 1.4) | 0.65 (0.47 - 0.91) | 0.47 (0.34 - 0.66) | 0.31 (0.26 - 0.38) |
| On arrival UE | 1.2 (1.0 - 1.5) | 0.96 (0.75 - 1.2) | 0.57 (0.40 - 0.81) | 0.40 (0.28 - 0.59) | 0.25 (0.20 - 0.32) |
| On arrival enforced | 1.2 (0.96 - 1.4) | 0.93 (0.72 - 1.2) | 0.56 (0.39 - 0.80) | 0.39 (0.27 - 0.57) | 0.24 (0.20 - 0.31) |
| At end |  | 0.97 (0.75 - 1.2) | 0.55 (0.40 - 0.76) | 0.43 (0.32 - 0.58) | 0.33 (0.27 - 0.41) |
| All infections are symptomatic |  |  |  |  |  |
| None | 1.8 (1.5 - 2.2) | 1.2 (0.99 - 1.5) | 0.66 (0.49 - 0.87) | 0.49 (0.37 - 0.64) | 0.37 (0.30 - 0.44) |
| 3 days prior | 1.7 (1.5 - 2.1) | 1.2 (0.97 - 1.5) | 0.64 (0.48 - 0.85) | 0.47 (0.36 - 0.63) | 0.35 (0.29 - 0.43) |
| On arrival UE | 1.3 (1.1 - 1.5) | 0.98 (0.79 - 1.2) | 0.52 (0.38 - 0.71) | 0.38 (0.28 - 0.52) | 0.27 (0.22 - 0.32) |
| On arrival enforced | 1.3 (1.1 - 1.5) | 0.97 (0.78 - 1.2) | 0.52 (0.38 - 0.70) | 0.37 (0.28 - 0.51) | 0.26 (0.22 - 0.32) |
| At end |  | 0.99 (0.80 - 1.2) | 0.53 (0.40 - 0.70) | 0.43 (0.33 - 0.55) | 0.36 (0.30 - 0.44) |
| Base scenario |  |  |  |  |  |
| None | 1.7 (1.5 - 2.0) | 1.2 (1.0 - 1.4) | 0.67 (0.54 - 0.84) | 0.49 (0.40 - 0.63) | 0.35 (0.31 - 0.41) |
| 3 days prior | 1.7 (1.4 - 1.9) | 1.2 (1.00 - 1.4) | 0.65 (0.52 - 0.82) | 0.47 (0.39 - 0.61) | 0.34 (0.29 - 0.39) |
| On arrival UE | 1.3 (1.1 - 1.5) | 0.97 (0.82 - 1.2) | 0.54 (0.42 - 0.71) | 0.39 (0.30 - 0.52) | 0.26 (0.23 - 0.31) |
| On arrival enforced | 1.2 (1.1 - 1.4) | 0.95 (0.81 - 1.1) | 0.53 (0.42 - 0.71) | 0.38 (0.30 - 0.52) | 0.25 (0.22 - 0.31) |
| At end |  | 0.98 (0.83 - 1.2) | 0.54 (0.43 - 0.70) | 0.43 (0.35 - 0.55) | 0.35 (0.30 - 0.41) |
| Infected at time of arrival |  |  |  |  |  |
| None | 2.5 (2.1 - 2.9) | 2.3 (2.0 - 2.7) | 1.6 (1.3 - 1.9) | 1.1 (0.85 - 1.4) | 0.55 (0.45 - 0.69) |
| 3 days prior | 2.5 (2.1 - 2.9) | 2.3 (2.0 - 2.7) | 1.6 (1.3 - 1.9) | 1.1 (0.85 - 1.4) | 0.55 (0.45 - 0.69) |
| On arrival UE | 2.5 (2.1 - 2.9) | 2.3 (2.0 - 2.7) | 1.6 (1.3 - 1.9) | 1.1 (0.85 - 1.4) | 0.55 (0.45 - 0.69) |
| On arrival enforced | 2.5 (2.1 - 2.9) | 2.3 (2.0 - 2.7) | 1.6 (1.3 - 1.9) | 1.1 (0.85 - 1.4) | 0.55 (0.45 - 0.69) |
| At end |  | 2.1 (1.8 - 2.4) | 1.3 (1.0 - 1.6) | 0.89 (0.67 - 1.2) | 0.53 (0.44 - 0.65) |
| Low compliance (50% for quarantine, with symptoms, or with a positive test; 70% for both symptoms and a positive test) |  |  |  |  |  |
| None | 2.3 (2.0 - 2.7) | 1.9 (1.7 - 2.3) | 1.5 (1.2 - 1.8) | 1.3 (1.1 - 1.5) | 1.2 (1.0 - 1.4) |
| 3 days prior | 2.2 (1.9 - 2.5) | 1.9 (1.6 - 2.1) | 1.4 (1.2 - 1.7) | 1.2 (1.1 - 1.5) | 1.1 (0.96 - 1.3) |
| On arrival UE | 2.0 (1.7 - 2.3) | 1.7 (1.5 - 2.0) | 1.3 (1.1 - 1.5) | 1.1 (0.96 - 1.3) | 1.0 (0.87 - 1.2) |
| On arrival enforced | 1.6 (1.4 - 1.8) | 1.4 (1.2 - 1.7) | 1.1 (0.90 - 1.3) | 0.94 (0.79 - 1.1) | 0.81 (0.71 - 0.93) |
| At end |  | 1.8 (1.6 - 2.1) | 1.4 (1.2 - 1.7) | 1.3 (1.1 - 1.5) | 1.2 (1.0 - 1.3) |
| Perfect compliance (100% compliance for quarantine and isolation) |  |  |  |  |  |
| None | 1.4 (1.1 - 1.6) | 0.78 (0.63 - 0.96) | 0.25 (0.15 - 0.39) | 0.11 (0.048 - 0.21) | 0.004 (0.000 - 0.020) |
| 3 days prior | 1.3 (1.1 - 1.5) | 0.76 (0.62 - 0.94) | 0.25 (0.15 - 0.39) | 0.11 (0.047 - 0.20) | 0.004 (0.000 - 0.020) |
| On arrival UE | 0.99 (0.84 - 1.1) | 0.66 (0.53 - 0.82) | 0.23 (0.14 - 0.37) | 0.10 (0.045 - 0.20) | 0.004 (0.000 - 0.020) |
| On arrival enforced | 0.99 (0.84 - 1.1) | 0.66 (0.53 - 0.82) | 0.23 (0.14 - 0.37) | 0.10 (0.045 - 0.20) | 0.004 (0.000 - 0.020) |
| At end |  | 0.58 (0.46 - 0.74) | 0.16 (0.084 - 0.28) | 0.063 (0.023 - 0.14) | 0.002 (0.000 - 0.014) |
| Travelers are equally likely to be in any disease phase (including symptomatic) |  |  |  |  |  |
| None | 1.5 (1.3 - 1.7) | 1.0 (0.88 - 1.2) | 0.57 (0.46 - 0.74) | 0.42 (0.34 - 0.55) | 0.31 (0.26 - 0.36) |
| 3 days prior | 1.4 (1.2 - 1.6) | 0.99 (0.85 - 1.2) | 0.55 (0.43 - 0.70) | 0.40 (0.32 - 0.52) | 0.29 (0.25 - 0.33) |
| On arrival UE | 1.1 (0.96 - 1.2) | 0.83 (0.69 - 0.99) | 0.46 (0.35 - 0.61) | 0.33 (0.25 - 0.45) | 0.22 (0.19 - 0.27) |
| On arrival enforced | 1.1 (0.93 - 1.2) | 0.81 (0.68 - 0.98) | 0.45 (0.34 - 0.60) | 0.32 (0.25 - 0.44) | 0.22 (0.19 - 0.26) |
| At end |  | 0.84 (0.71 - 1.0) | 0.47 (0.37 - 0.61) | 0.37 (0.30 - 0.48) | 0.31 (0.26 - 0.35) |
| Travelers have not developed symptoms before arriving |  |  |  |  |  |
| None | 1.9 (1.6 - 2.2) | 1.3 (1.1 - 1.6) | 0.72 (0.58 - 0.91) | 0.53 (0.43 - 0.68) | 0.38 (0.33 - 0.45) |
| 3 days prior | 1.8 (1.6 - 2.0) | 1.3 (1.1 - 1.5) | 0.70 (0.57 - 0.88) | 0.51 (0.42 - 0.66) | 0.37 (0.32 - 0.42) |
| On arrival UE | 1.4 (1.2 - 1.6) | 1.1 (0.91 - 1.3) | 0.59 (0.46 - 0.76) | 0.42 (0.33 - 0.56) | 0.29 (0.25 - 0.34) |
| On arrival enforced | 1.4 (1.2 - 1.5) | 1.0 (0.89 - 1.2) | 0.58 (0.46 - 0.75) | 0.41 (0.33 - 0.55) | 0.28 (0.24 - 0.33) |
| At end |  | 1.1 (0.91 - 1.2) | 0.58 (0.47 - 0.74) | 0.46 (0.38 - 0.58) | 0.38 (0.33 - 0.44) |

## SUPPLEMENTAL FIGURES

**Figure S1:**
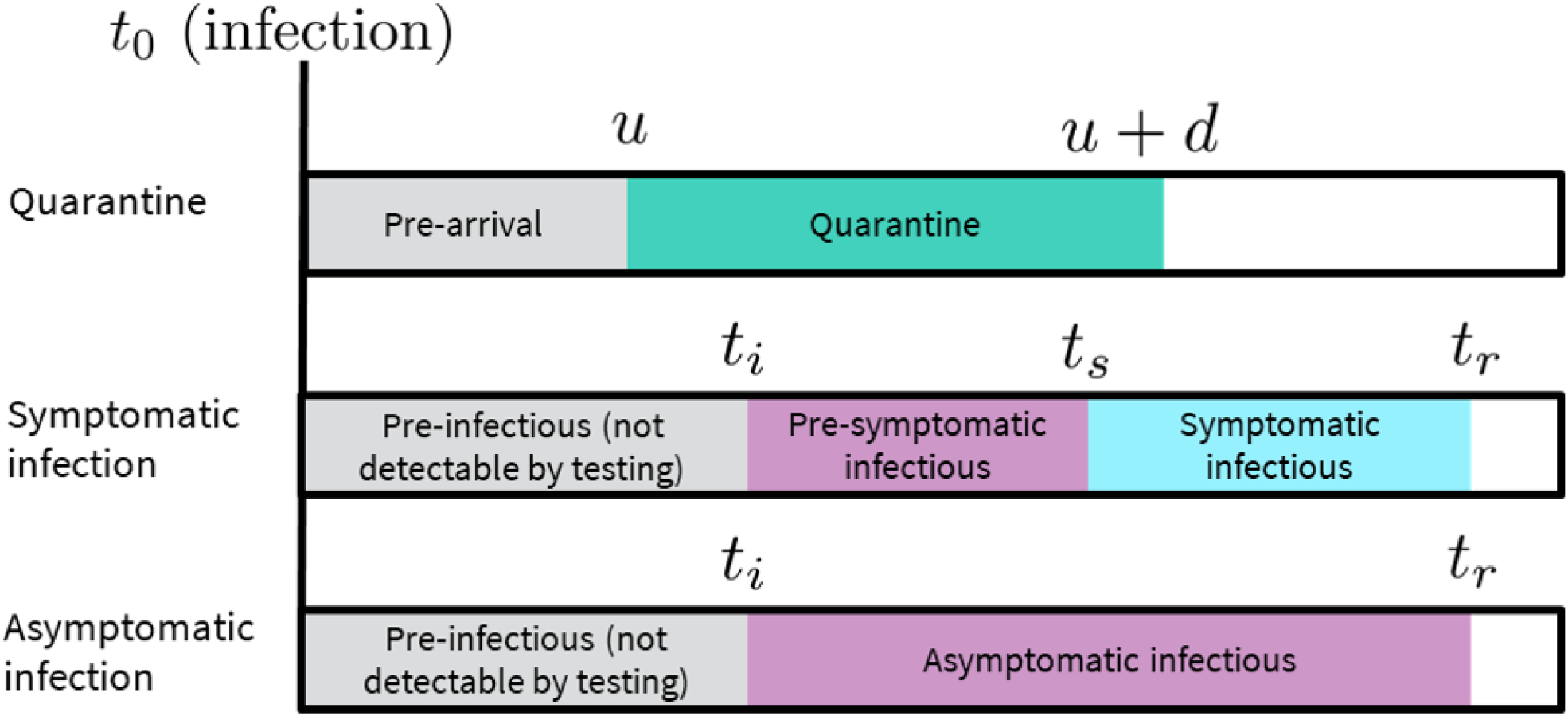
Schematic of quarantine and infection phases as represented in the model. t_0_ is the time of infection; ti is the time infectiousness begins, tr is the time individuals recover from being infectious, and ts is the time of symptom onset for those with symptomatic infections. u is the time of arrival and d is the duration of quarantine. Community transmission risk can occur during the mandatory quarantine period for non-compliant travelers or after quarantine for travelers who do not recover before quarantine end. We assume most travelers self-isolate for the duration of the infectious phase after symptom onset or receiving a positive test.

**Figure S2:**
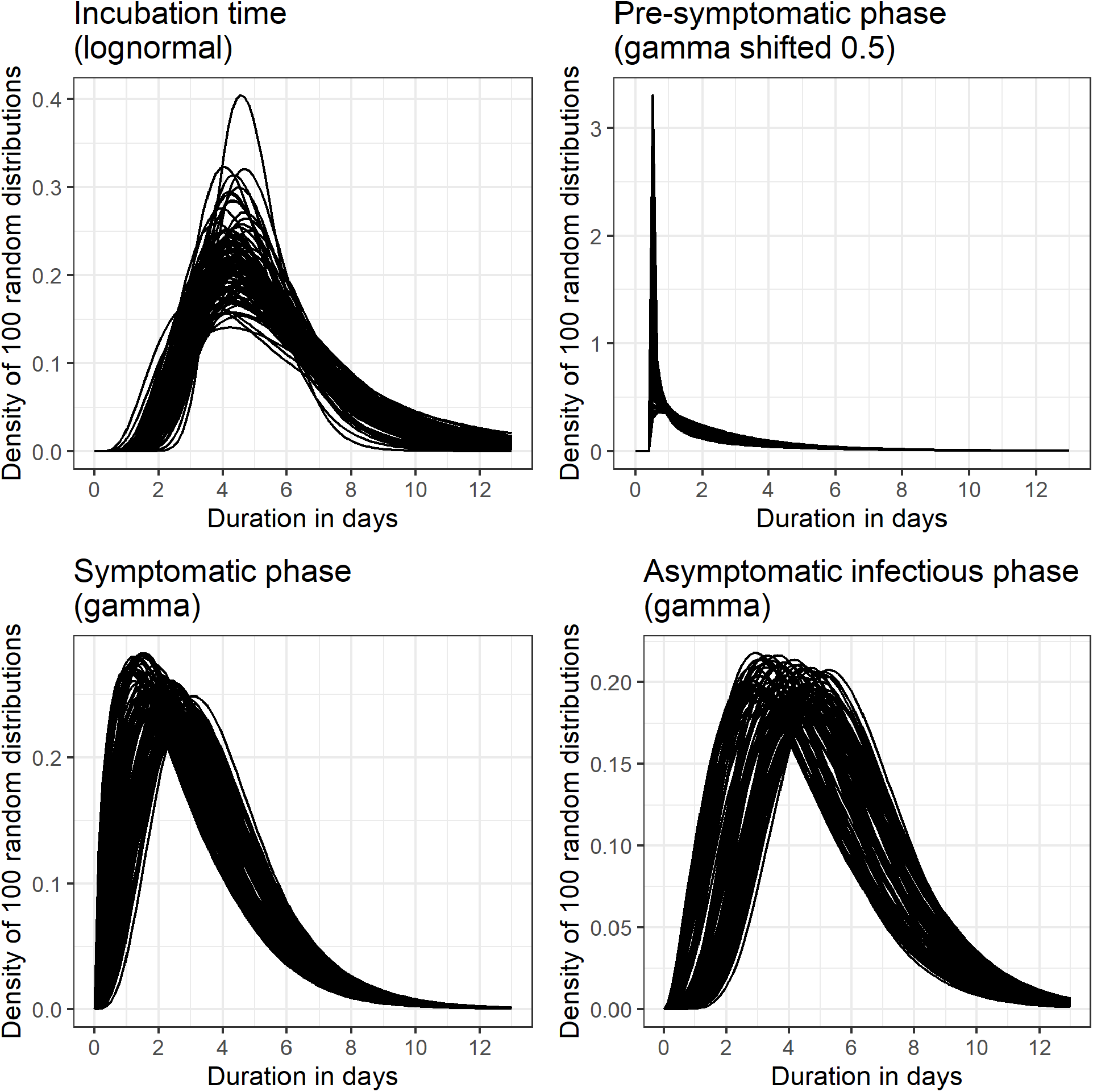
Sample of 100 distributions for durations. Incubation time (top-left) generated using code from Lauer 2020 [7] that generates bootstrapped posterior distributions from a parametric accelerated failure time model calibration. Other distributions generated by varying the mean and variance of distributions from Moghadas 2020 [8] by ±20% and converting to gamma distribution parameters using the relationships μ = ab and σ^2^ = ab^2^ where μ and σ are the mean and variance of the gamma distribution and a and b are the shape and scale parameters. For both symptomatic and asymptomatic infections, duration of the pre-infectious-infectious phase is calculated by subtracting the pre-symptomatic phase duration from the incubation time. If the calculated start of the pre-symptomatic-infectious phase is less than 0.5 days from infection, 0.5 days is used instead.

**Figure S3:**
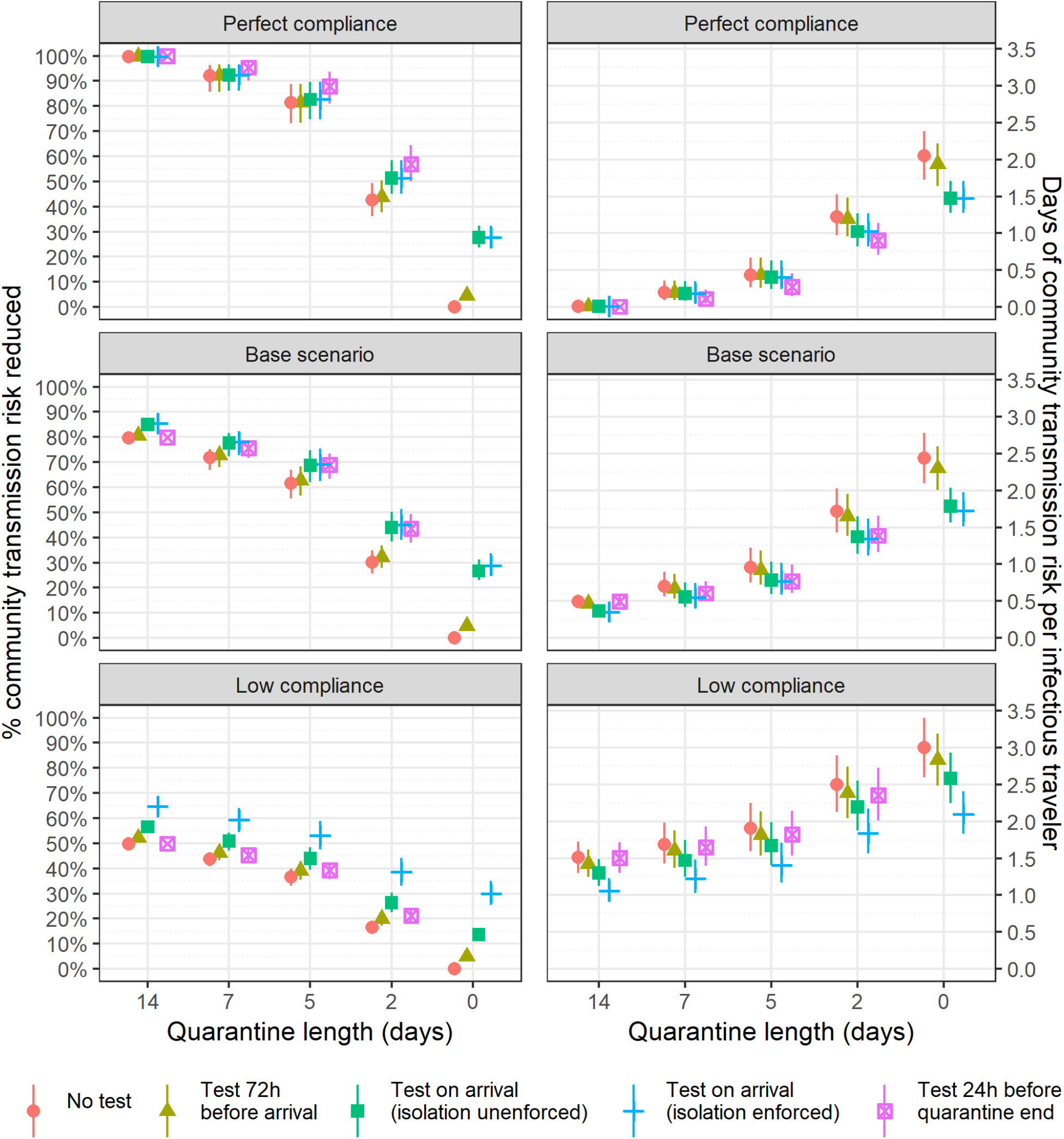
Estimated percent community transmission risk reduced (left) and days of community transmission risk per infected traveler (right) for each quarantine and testing policy in three scenarios with different levels of quarantine and isolation compliance.

**Figure S4:**
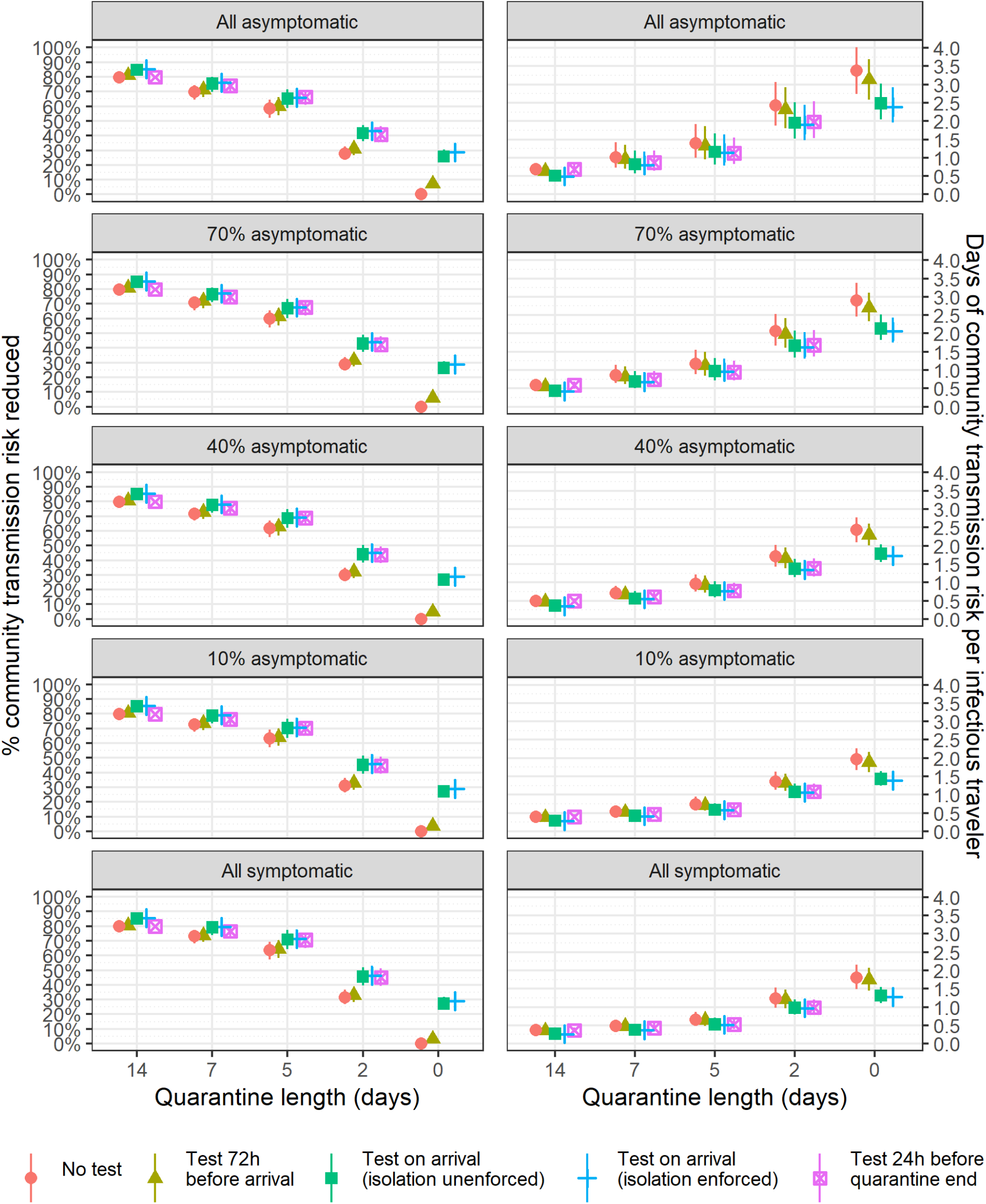
Estimated percent community transmission risk reduced (left) and days of community transmission risk per infected traveler (right) for each quarantine and testing policy in five scenarios with different levels of asymptomatic infection.

**Figure S5:**
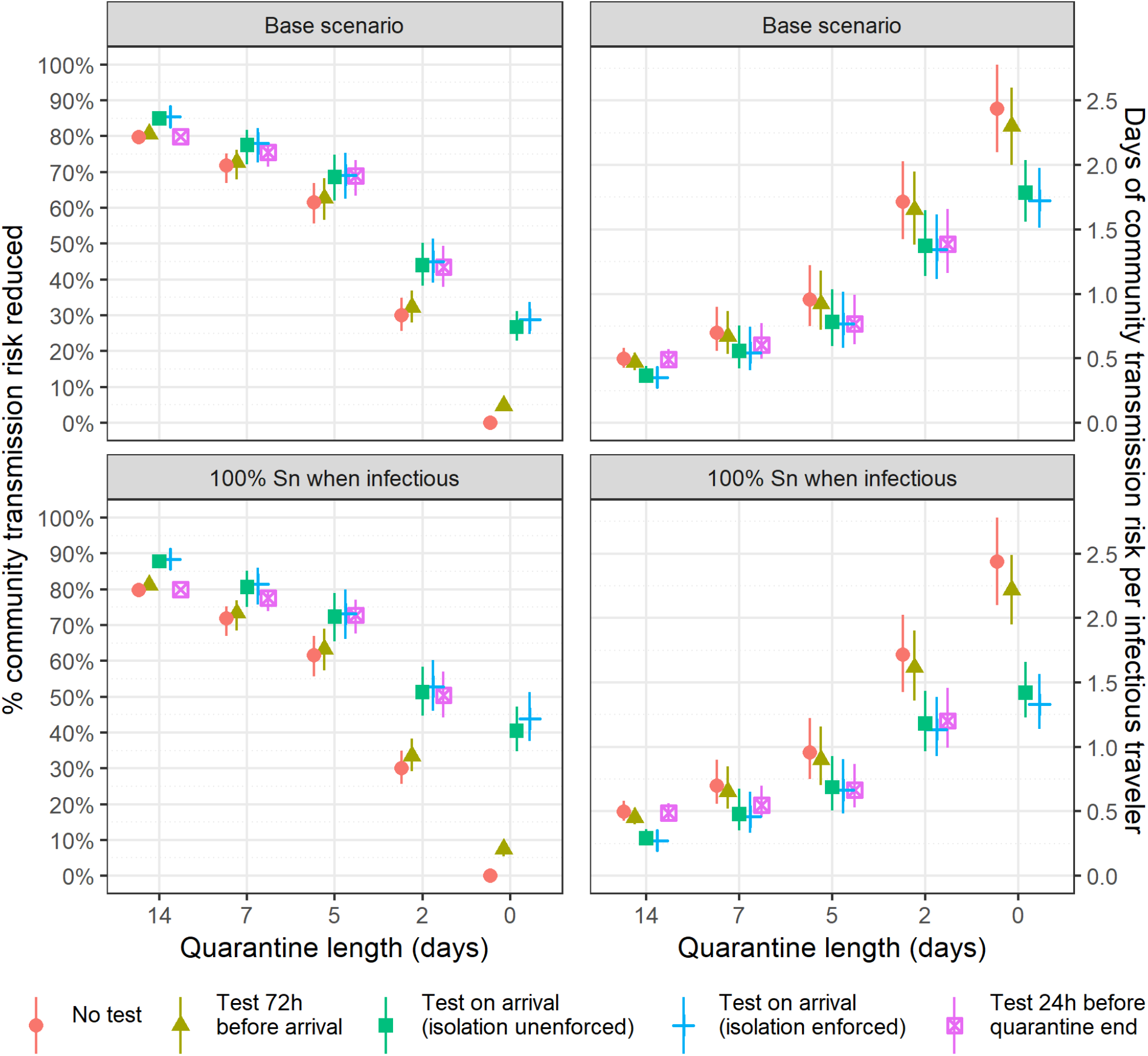
Estimated percent community transmission risk reduced (left) and days of community transmission risk per infected traveler (right) for each quarantine and testing policy for a test with 100% sensitivity for all travelers in an infectious phase compared to base case assumption (70% sensitivity for symptomatic infections; 60% sensitivity for asymptomatic infections).

**Figure S6:**
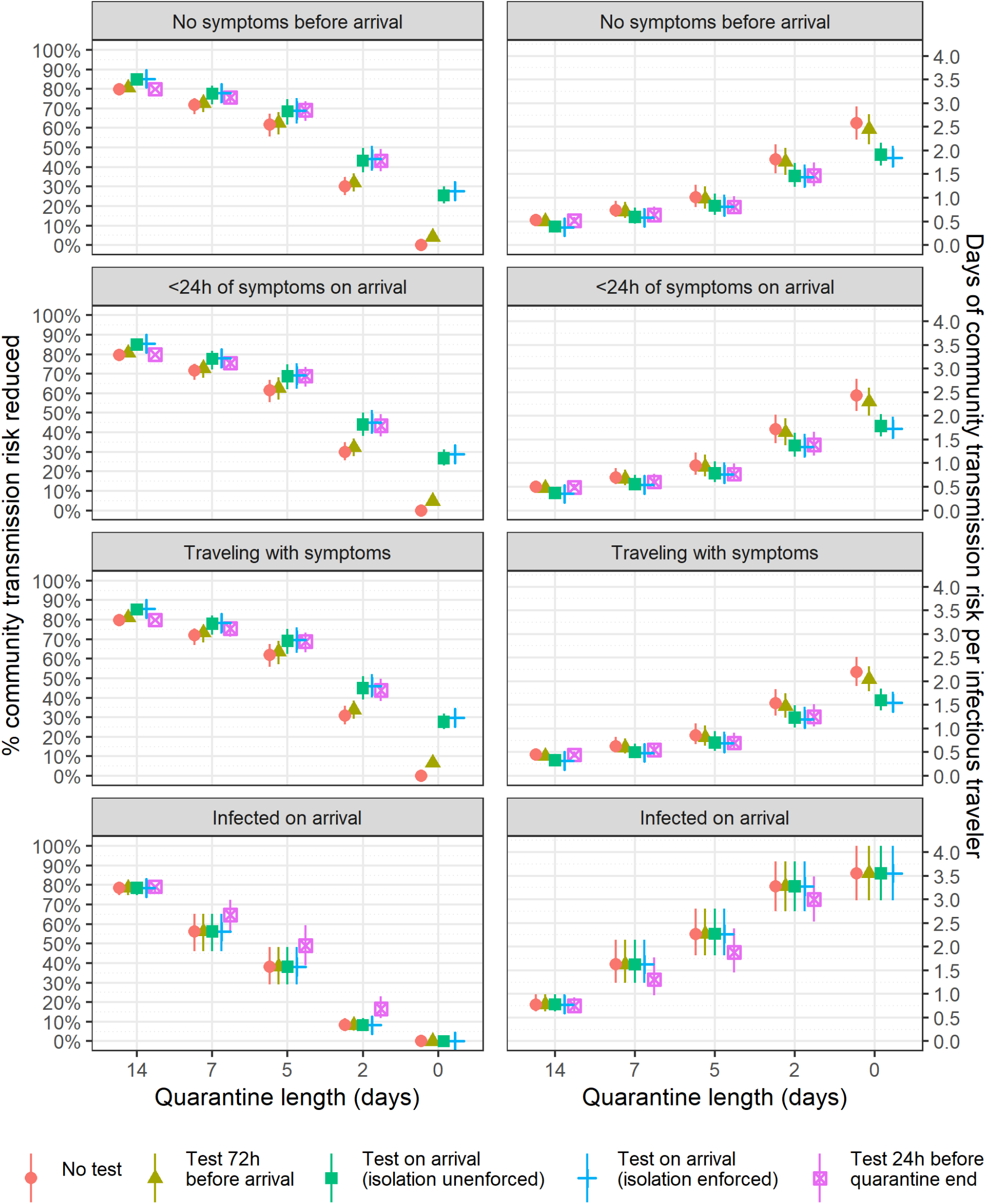
Estimated percent community transmission risk reduced (left) and days of community transmission risk per infected traveler (right) for each quarantine and testing policy in four scenarios with different assumptions about the distribution of the timing of infections relative to infected travelers’ arrival time.

**Figure S7:**
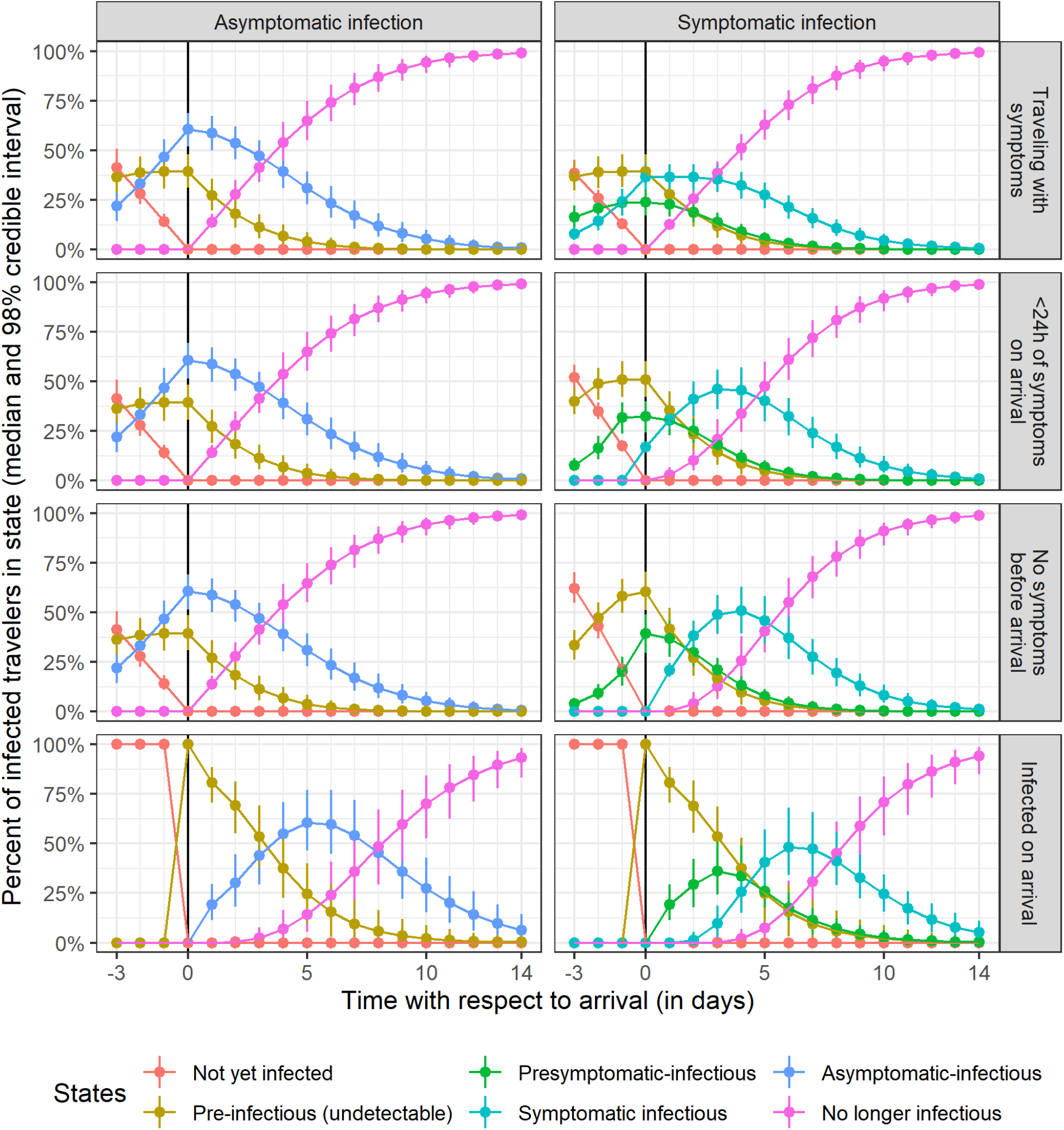
Distribution of infection state membership for infected travelers with asymptomatic (left) or symptomatic (right) infection before and after arrival under four different infection timing assumptions. Travelers cannot yet test positive in the ‘not yet infected’ or ‘pre-infectious’ states.

## REFERENCES

1. Shi Q, Hu Y, Peng B, et al. Effective control of SARS-CoV-2 transmission in Wanzhou, China. Nature Medicine. 2021;27(1):86–93. doi:10.1038/s41591-020-01178-5

2. Bisoffi Z, Pomari E, Deiana M, et al. Sensitivity, specificity and predictive values of molecular and serological tests for COVID-19. A longitudinal study in emergency room. medRxiv. 2020. doi:10.1101/2020.08.09.20171355

3. Russell WA, Buckeridge DL. Code and data repository for modeling effectiveness of quarantine and testing to prevent COVID-19 transmission from arriving travelers. Zenodo. 2020. doi:10.5281/zenodo.4107124

4. Wells CR, Townsend JP, Pandey A, et al. Optimal COVID-19 quarantine and testing strategies. Nature Communications. 2021;12(1):1–9. doi:10.1038/s41467-020-20742-8

5. Ashcroft P, Lehtinen S, Angst DC, Low N, Bonhoeffer S. Quantifying the impact of quarantine duration on COVID-19 transmission. 2020;10. doi:10.1101/2020.09.24.20201061

6. Quilty BJ, Clifford S, Hellewell J, et al. Quarantine and testing strategies in contact tracing for SARS-CoV-2: a modelling study. The Lancet Public Health. 2021;0(0). doi:10.1016/s2468-2667(20)30308-x

7. Lauer SA, Grantz KH, Bi Q, et al. The incubation period of coronavirus disease 2019 (CoVID-19) from publicly reported confirmed cases: Estimation and application. Annals of Internal Medicine. 2020;172(9):577–582. doi:10.7326/M20-0504

8. Moghadas SM, Fitzpatrick MC, Sah P, et al. The implications of silent transmission for the control of COVID-19 outbreaks. Proceedings of the National Academy of Sciences. 2020;117(30):17513–17515. doi:10.1073/pnas.2008373117

